# How do environmental, economic and health factors influence regional vulnerability to COVID-19?

**DOI:** 10.1101/2020.04.09.20059659

**Authors:** Pejman Tahmasebi, Salome M.S. Shokri-Kuehni, Muhammad Sahimi, Nima Shokri

## Abstract

We have studied the correlations between twelve *environmental, economic* and *health* variables, by carrying out a statistical analysis of the fatality rate of COVID-19 in 14 countries. Our statistical analysis indicates that, among the 12 variables, the diabetes percentage of the total population and the extent of the population ages 65 and older in each country are correlated most strongly with the total number of deaths in them. Although the strength of the correlations between the variables and the total ND may change as the ongoing pandemic evolves, the study highlights the importance of integrating regional-specific variables in the modelling efforts aimed at projecting how the spread of the virus may influence different parts of the world.

## 1. Introduction

As of 7 April, 2020, there are 1,279,722 confirmed cases of Coronavirus disease (COVID-19) and 72,616 confirmed deaths due to the ongoing COVID-19 pandemic affecting 211 countries, areas or territories globally.^1^ This unfolding situation has caused unprecedented conditions around the globe, affecting not only human health, but also the global economic, international trade and relations, as well as friction. The extremely challenging global state of affairs has also motivated many experts to investigate various aspects related to the fast spreading virus.^2–6^ To this end, several groups have been trying to model the spread of this virus under different scenarios, aiming to project the possible lives that may be lost due to COVID-19, including the influential works at Imperial College London^7^ and University of Washington.^8^

One of the most important factors in all the models used to describe any infectious disease outbreak is its reproduction number *R*_0_. When *R*_0_ > 1, the infection is statistically capable of spreading in a population, whereas *R*_0_ ≤ 1 indicates breakage of the chain of the spread of the virus, which eventually ends the transmission.^4^ What is particularly concerning about COVID-19 is its current reproduction number in the range 2.5 - 2.9, which may change amid the evolving pandemic.^4^ To put the current *R*_0_ value of COVID-19 into perspective, one can compare it with the estimated reproduction rate of 1.7–2.0 for the first wave of the 1918 pandemic^9^ (the so-called Spanish flu), which killed about 50 million people worldwide.

Such a high reproduction number triggered implementation of a range of intensive non-pharmaceutical interventions in many countries around the world as an attempt to control and suppress the spread of COVID-19. Such measures include, but not limited to, school closures, banning public gathering, social distancing, and lockdown at various scales, ordered by the authorities in order to slow down the spread of the virus. Mathematical models^7^ are used to quantify how the implementation of such interventions can possibly slow the spread of COVID-19 or “flatten the curve”. Valuable insights are obtained by such modelling, which can form an important part of policymaking for helping the government actions to alter the course of the outbreak.

A special emphasis has been placed on social distancing, while other regional-scale and inherent variables have been ignored. As such, the current modelling efforts are focused mainly on how distinct social distancing scenarios and government intervention may modify the spread of COVID-19, with less attention paid to the possible impact of the pre-existing regional environmental, economic, and health variables, which may influence the vulnerability to COVID-19. In other words, drawing a more realistic picture about the spread and fatality rate of COVID-19 requires consideration of a combination of such parameters, as the rate of fatality is not all about the actions that are taken, but also due to many other factors developed in each region over many years due to a variety of environmental, health, culture, economic and political reasons that we might have little or no control on in the limited time that the world has to deal with COVID-19.

In fact, many questions regarding the spread and fatality of COVID-19 have remained unanswered. For example, when similar government interventions are implemented, why different regions experience different fatality rates? How and which environmental, cultural, economic, and health variables define the vulnerability of a country to COVID-19? In this study, we present a different view on the problem by classifying them into three groups that we explain shortly. To do so, we have studied the *correlations* between twelve *environmental, economic* and *health* variables, and have carried out a statistical analysis of the fatality rate of COVID-19 in many countries, as a first step for highlighting how country-specific variables may influence the fatality rate. Such a study should contribute toward developing realistic models for describing the fatality rate of the Coronavirus, other than simply considering parameters whose influence may be minor.

## 2. Methods

We considered 14 countries whose statistics have been available to the public from the emergence of COVID-19. They represent the total number of deaths (ND) as of 06 April 2020 according to the daily real-time death data released by the World Health Organization^1^. These countries include Italy (IT), Spain (ES), China (CN), the U.S., France (FR), Iran (IR), the UK, Netherlands (NL), Germany (DE), Belgium (BE), Switzerland (CH), South Korea (KR), Brazil (BR), and Sweden (SE). The correlation between the aforementioned 12 different variables with the total ND in each country was investigated. The variables include prevalence of obesity among adults, diabetes, cancer, smoking, insufficient physical activity among adults aged 18+ years, high blood pressure, high total cholesterol among adults aged 25+ year, population aged 65 year and older, population aged 15-64 year, CO_2_ per capita emission, gross domestic product, and gross national income.

In our analysis, we used a linear model to study the interactions between the aforementioned variables. The model describes general relationships between the variables and the target variable, the total ND. We used a least-squares method to fit the model and identify the variables, which have significant correlation with the total ND by defining their coefficients. The general form of the model is given by

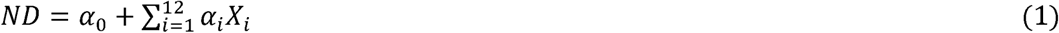

where *ND* represents the total number of deaths, *α*_0_ is a constant (equal to 1), *α*_*i*_ are the fitting coefficients, and *X*_*i*_ are the environmental, economic and health variables considered in the present investigation. To identify the most accurate equation, we used a stepwise algorithm, whereby systematic adding and removing the variables from Eq. (1) in the presence of all the variables were carried out. The method keeps/removes the variables based on their importance concerning the target variable, i.e., ND. Thus, it starts with a large initial model, i.e., one with a large number of variables, and then compares the explanatory power of incrementally larger and smaller models. More specifically, a forward--backward model is used to determine the final equation. It uses p-value and t-statistic to verify how a model behaves with and without a variable. As such, a variable is assigned a zero coefficient if it is not already in the model. If the remaining variables have p-values less than a threshold, the variable with the smallest p-value is added to the model. Otherwise, if the existing variables in the model have p-values larger than the threshold, the variable with the largest p-value is removed. Finally, the model is terminated if no improvements are observed.

## 3. Results and Discussions

The data provided by World Health Organization (WHO)^10^, the World Bank^11^, and the Emission Database for Global Atmospheric Research (EDGAR) of the European Commission^12^ were used to obtain the 12 environmental, economic and heath variables investigated in this study, with the data presented in Figure 1 (a,b). Moreover, we used the daily death data in each country released by the WHO^13^ with the data presented in Fig. 1 (c) and provided the general trends for ND.

**Fig. 1.**
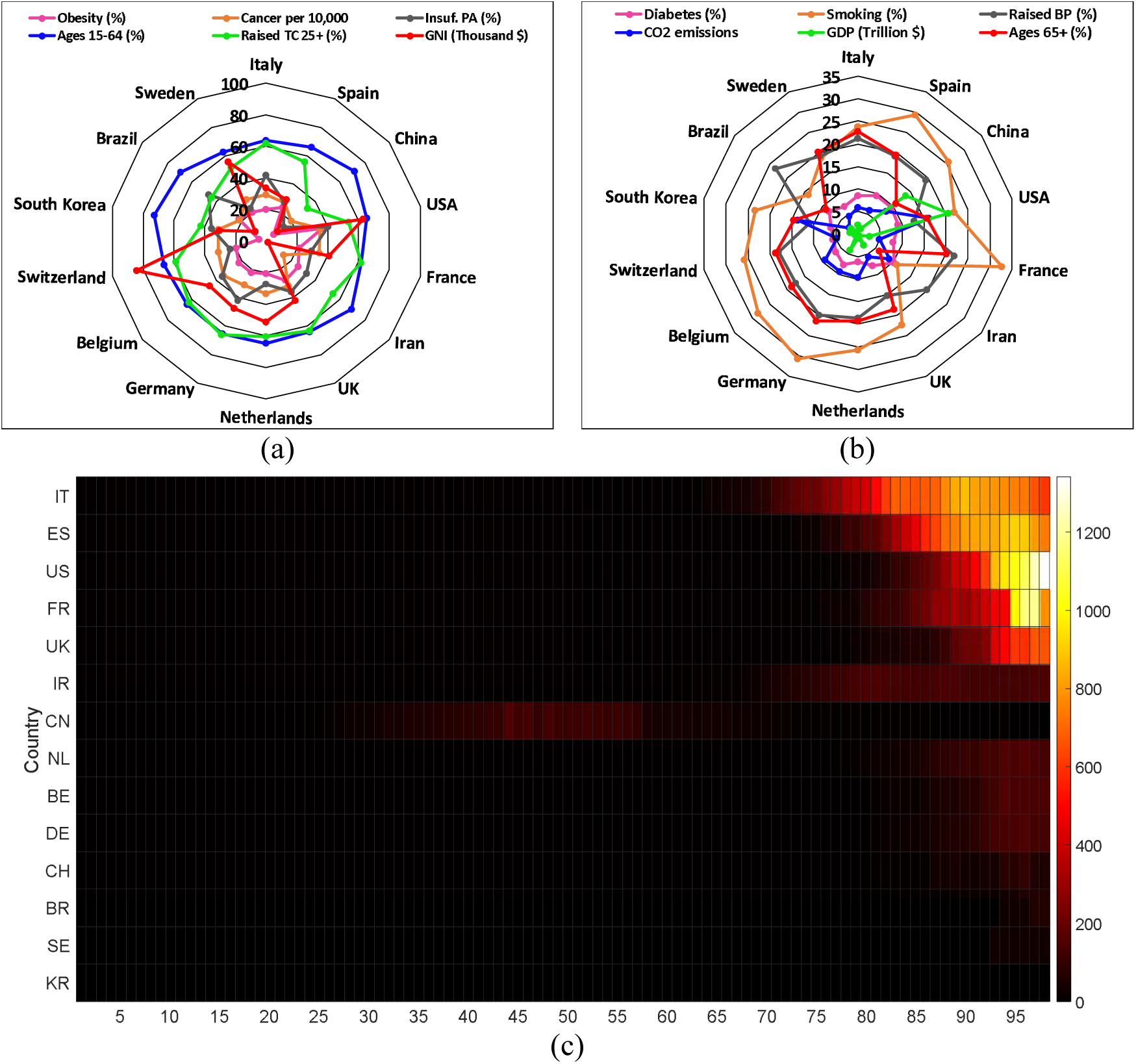
Top row (a, b): Prevalence of obesity among adults, BMI ≥ 30, age-standardized in 2016 labelled as “Obesity (%),” cancer data age-standardized incidence rates per 10,000 labelled as “Cancer per 10,000,” prevalence of insufficient physical activity among adults aged 18+ years in 2016 labelled as “Ins. PA (%),” population aged 15-64 labelled as “Ages 15-64 (%),” high total cholesterol (>= 5.0 mmol/L) (age-standardized estimate) in 2008 labelled as “Raised TC 25+,” gross national income in 2018 labelled as “GNI (in $thousands),” diabetes % of total population in 2016 labelled as “Diabetes (%),” prevalence of smoking in 2016 labelled as “Smoking (%),” raised blood pressure (SBP ≥ 140 or DBP ≥ 90), age-standardized (%) labelled as “Raised BP (%),” CO_2_ per capita emissions in 2018 labelled as “CO_2_ emission,” gross domestic product in 2018 labelled as “GDP (in $trillions)”, and population aged 65 and older labelled as “Ages 65+” in each country. The data for obesity, diabetes, raised blood pressure, raised total cholesterol, insufficient physical activity are from the World Health Organization, those for age population, prevalence of smoking, GDP and GNI from the World Bank, while the data for CO_2_ per capita emissions are from Emission Database for Global Atmospheric Research (EDGAR) of the European Commission. (c) The daily death due to COVID-19 in each country. The colors represent the 3-day average total number of deaths.

The data presented in Fig. 1 enables us to investigate the simultaneous relation between the 12 variables considered in the present study, on the one hand, and the total ND, on the other hand, whereas the relationship between the variables compared to other countries are summarized. Similarly, we also have analysed the trends of each variable and the ND separately in order to provide how each variable affects the ND. The results are presented in Figure 2.

**Fig. 2.**
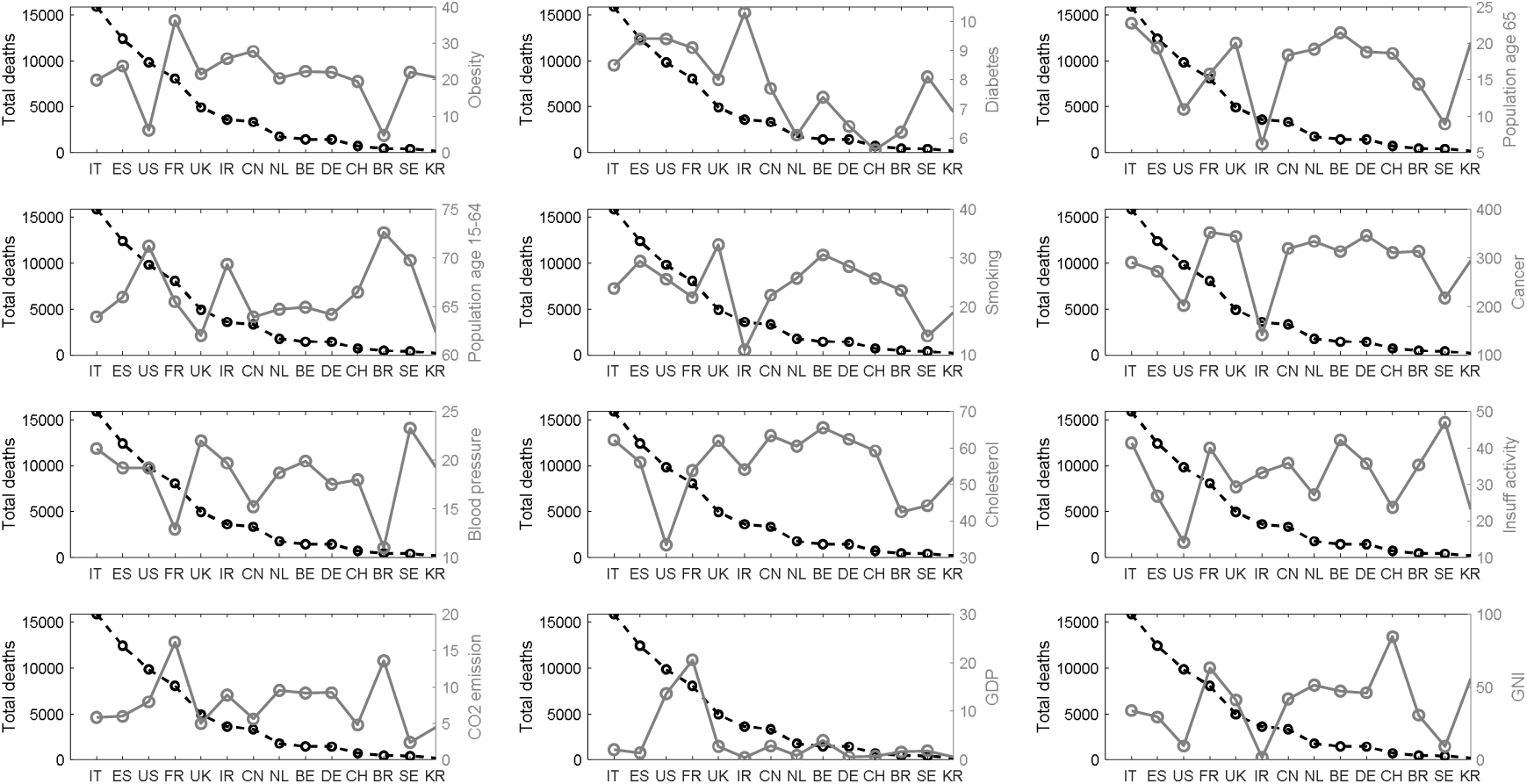
The relation between the total ND (as of 06 April 2020) and the 12 environmental, economic and health variables considered in the analysis. The variables are the same as those explained in the caption of Fig. 1.

To identify quantitatively the variables that are correlated with the total ND recorded in each country, the statistical procedure explained in the previous section was employed. Our results show that among the variables considered in the present study, the diabetes percentage of the total population and the population age 65 and higher had significant correlation with the total ND recorded in each country as of 06 April 2020. While the effect of the latter factor has been known, the fact that our statistical model correctly highlights its correlation indicates its internal consistency. The estimated coefficients for these two variables are summarized in Table 1.

**Table 1.**
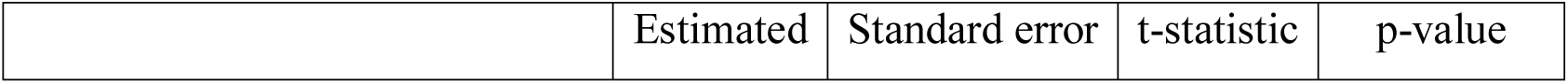
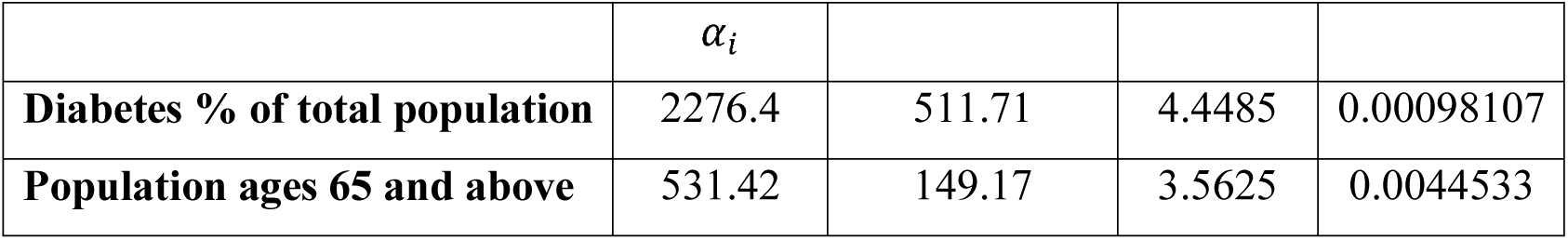
A summary of the extracted statistics for the implemented model

|  | Estimated | Standard error | t-statistic | p-value |
| --- | --- | --- | --- | --- |

| | $\alpha_i$ | | | |
| --- | --- | --- | --- | --- |
| **Diabetes % of total population** | 2276.4 | 511.71 | 4.4485 | 0.00098107 |
| **Population ages 65 and above** | 531.42 | 149.17 | 3.5625 | 0.0044533 |

To further demonstrate the correlation of our findings with the total ND, the spatial relationship between the total ND, diabetes percentage of the total population and the population ages 65 and higher is shown in Figure 3, where the colour map presents the total ND in each country. Figure 3 suggests that the countries with a higher percentage of diabetes and population ages 65 and older experience more fatality due to COVID-19, at least as of 06 April 2020.

**Fig. 3.**
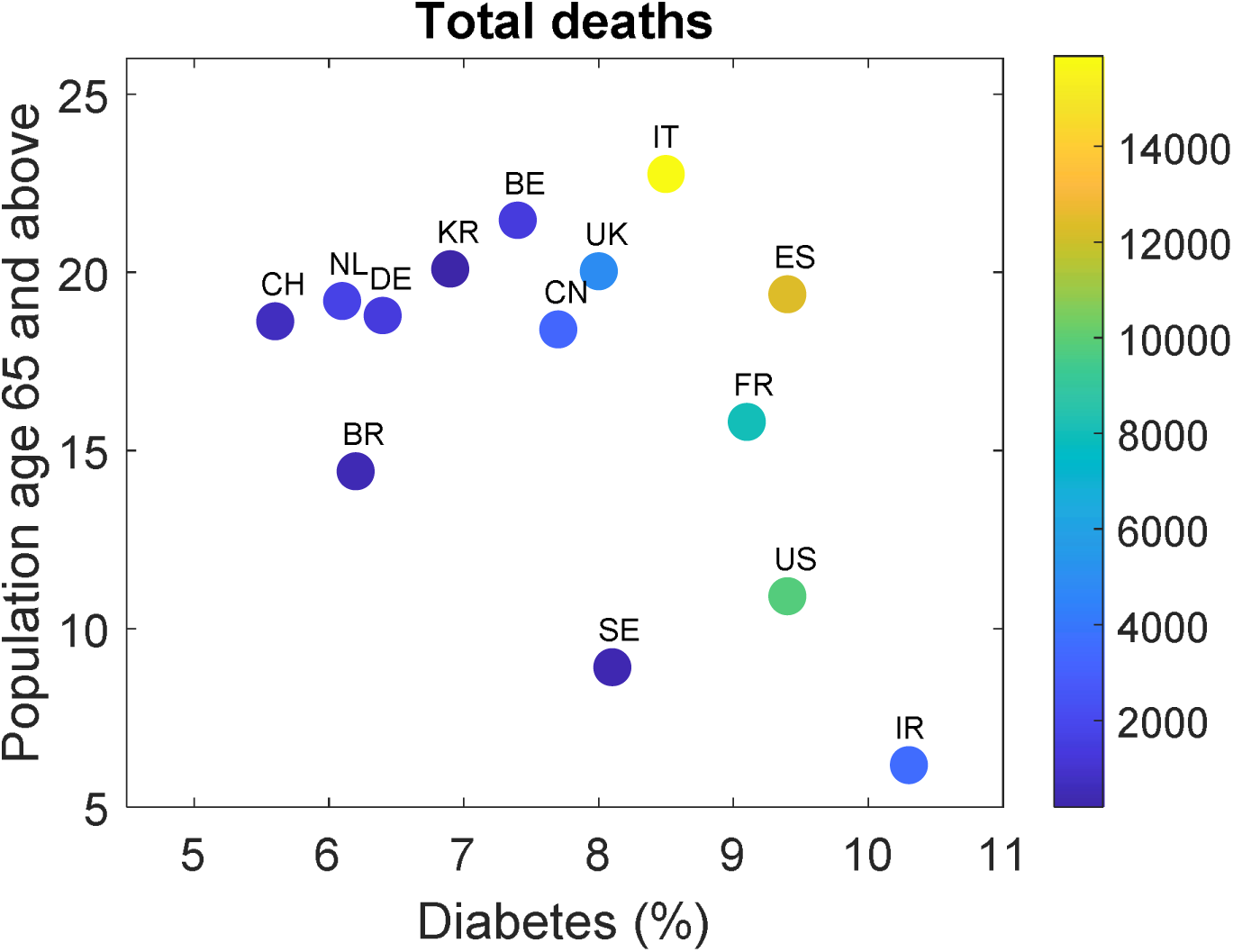
The spatial relation between the diabetes percentage of the total population, the population ages 65 and higher, and the total number of deaths ND, recorded in each country due to COVID-19, as of 06 April 2020. The colour map indicates the total number of death (ND).

## 4. Summary and Conclusions

We have investigated the correlation between 12 different environmental, economic and health variables with the total number of deaths in 14 countries, caused by COVID-19, as of 06 April 2020. Our statistical analysis revealed that, among the investigated variables, the diabetes percentage of the total population and the population ages 65 and older in each country are correlated most strongly with the total number of deaths in each country.

Although the strength of the identified correlations between these variables and the total ND may change as the ongoing pandemic evolves, our study highlights the importance of integrating country-specific variables in the modelling efforts aimed at projecting how the spread of the novel virus may influence different parts of the world. Applying similar social-distancing measures in different regions of the world may have different effectiveness because of the effects of other variables that influence the vulnerability of a specific region or country to COVID-19. Identifying the economic, environmental, health and social variables affecting the vulnerability of people in different regions to this devastating virus is important to devising appropriate and thoughtful response for addressing the pandemic.

## Data Availability

The data used in this analysis are publically available with the corresponding links given in the manuscript.

## Authors’ contributions

Authors contributed equally to the paper.

## Conflict of interest statements

The authors declare no conflict of interests.

## Role of funding source

None

## Ethics committee approval

Not applicable

## Notes

### Competing Interest Statement

The authors have declared no competing interest.

